# Early-evening indoor and outdoor foraging by major malaria vectors in Nchelenge, Zambia

**DOI:** 10.1101/2025.09.27.25336603

**Authors:** Mary E. Gebhardt, Hannah L. Markle, Rachel S. Krizek, David Mbewe, Francis Kapasa Mulenga, James Sichivula Lupiya, Erin E. Barnett, Mbanga Muleba, Matthew M. Ippolito, Jennifer C. Stevenson, William J. Moss, Douglas E. Norris, the Southern Africa International Centers of Excellence for Malaria Research

**Affiliations:** The W. Harry Feinstone Department of Molecular Microbiology and Immunology, Johns Hopkins Bloomberg School of Public Health, Baltimore, Maryland, USA; Johns Hopkins Malaria Research Institute, Johns Hopkins Bloomberg School of Public Health, Baltimore, Maryland, USA; National Health Research & Training Institute, Ndola, Zambia; Department of Medicine, Johns Hopkins School of Medicine, Baltimore, Maryland, USA; Vector Biology Unit, Department of Epidemiology and Public Health, Swiss Tropical & Public Health Institute, Basel, Switzerland; Vector Control Product Testing Unit, Ifakara Health Institute, Bagamoyo, Tanzania; Department of Epidemiology, Johns Hopkins Bloomberg School of Public Health, Baltimore, Maryland, USA

**Author notes:** These authors contributed equally to this work (Mary E. Gebhardt, Hannah L. Markle). Information of the Southern Africa International Centers of Excellence for Malaria Research can be found online at https://www.niaid.nih.gov/research/icemr-regional-centers.

## Abstract

Despite a decade of indoor residual spraying, malaria remains holoendemic in Nchelenge District in northern Zambia. This study examined the hypothesis that the intransigence of malaria in Nchelenge District is due in part to early evening and outdoor foraging of local anopheline species. Three replicate collections were performed at 24 households using CDC light traps over 6 weeks during the dry season of 2019. Traps were set indoors, in animal pens, and in outdoor gathering spaces from 16:00-22:00, and overnight indoors in all households during the third replicate. Twelve anopheline species were collected in early-evening traps. The major vector, *Anopheles funestus,* dominated collections (n = 1,865, 68.2%) and were captured primarily indoors (n = 1,370, 73.4%). Specimens with detectable human blood and *Plasmodium falciparum* positive specimens were found in all three trap types, indicating that infected anopheline foraging occurred before 22:00. EIR estimates for early-evening anophelines varied spatially and by species and were highest among indoor caught *An. funestus* at inland locations (18.74 bites/person/6months). There was little difference in anopheline abundance, blooded rates, and sporozoite infection rate when evening and overnight traps were compared, indicating there may be similar anopheline activity before and after 22:00. Overall, this study adds to the growing body of evidence that malaria transmission is not limited to late-night, indoor settings in Nchelenge District, Zambia.

## Introduction

Despite a decade of annual indoor residual spraying (IRS), malaria transmission remains holoendemic in Nchelenge District, in northern Zambia. The well-recognized vectors in this area include *Anopheles funestus* sensu stricto (s.s.) and *An. gambiae* s.s., which are uniquely driven by local ecology and have different breeding preferences and spatial distributions across the district [1, 2]. Nchelenge District lies along the eastern banks of Lake Mweru, innervated by rivers, streams, and swampy areas. This ecology leads to elevated *An. funestus* populations during the dry season, likely due to decreased streamflow when precipitation is low or absent, while *An. gambiae* populations are present at relatively low abundance year-round, presumably taking advantage of ephemeral water bodies as their preferred breeding sites [1, 2]. Previous studies in Nchelenge District identified *An. funestus* as the dominant malaria vector, with entomologic inoculation rates (EIRs) ranging from 3.7-41.5 infectious bites/person/6 months (ib/p/6mo), while *An. gambiae* EIRs *range* from 0-5.9 infectious ib/p/6mo during the dry season [1].

Both *An. funestus* and *An. gambiae* are generally considered to be highly endophagic, endophilic, and anthropophilic across the African continent, yet IRS has had little impact on malaria rates or vector counts in Nchelenge District. The Southern and Central Africa International Center of Excellence for Malaria Research (ICEMR) performed impact evaluations of three years of IRS using Actellic® 300CS (Syngenta Group Company) and found a modest decrease in *An. funestus* populations but no impact on *An. gambiae* populations [3, 4]. Even a second round of IRS near the end of the rainy season had little impact on transmission intensity in this setting [5]. One explanation for these minimal temporary impacts is that anophelines are foraging both outdoors and indoors before people are protected by their bed nets.

There is growing concern for the selection of malaria vectors with behavior profiles that allow evasion of insecticide-treated bednets (ITNs) or IRS [6–10]. It has been hypothesized that IRS and ITNs select for these behaviors or result in species composition shifts, favoring species and/or populations with more plasticity in their foraging, biting, and resting behaviors [6, 11–13]. Lack of long-term longitudinal outdoor collections has made it difficult to demonstrate the extent of these behavioral changes, but a few studies have documented shifts in biting time or location [7, 14, 15] or shifts in species diversity over time [6, 11, 12]. In prior studies in Nchelenge District, most collections were performed indoors, with one small-scale study that collected anophelines outdoors targeting animal pens, places where people gather, and highly trafficked areas [16]. That study revealed a higher diversity of anophelines than previously recognized, raising the potential that outdoor foraging and malaria transmission risk was underestimated [16].

Understanding anopheline behavior and host-seeking preferences is essential for malaria control. Equally important is how human behavior and vector interactions influence the potential for malaria parasite transmission. It is valuable to take into consideration not only ITN use but other factors such as outdoor evening communal activities, cultural events, and outdoor sleeping habits that may leave people exposed to host-seeking anophelines [17, 18]. Documenting these human behaviors and linking them to anopheline biting times can yield insight into spatial and temporal risks for malaria transmission [17–20]. The study presented here assessed indoor and outdoor mosquito foraging and was performed to enhance our understanding of anophelines that bite in the early evening relative to overnight in Nchelenge District, Zambia and to assess spatial and temporal risk among participants during the dry season in 2019.

## Materials and Methods

### Study region

This study was performed from July to August 2019 in Nchelenge District, Zambia (**Fig 1A-C**). This district experiences three seasons: a rainy season from December to April, a cold dry season from May to August, and a hot dry season from September to November [1]. The malaria control program conducts annual IRS campaigns prior to the rainy season, usually in October. Due to high levels of pyrethroid resistance in both *An. funestus* and *An. gambiae,* IRS programs have used insecticides with various modes of action for IRS including Actellic® 300CS (Syngenta, Sweden) from 2013-2018 and Fludora® Fusion (Bayer Crop Science, Germany) from 2019-2020.

**Fig. 1.**
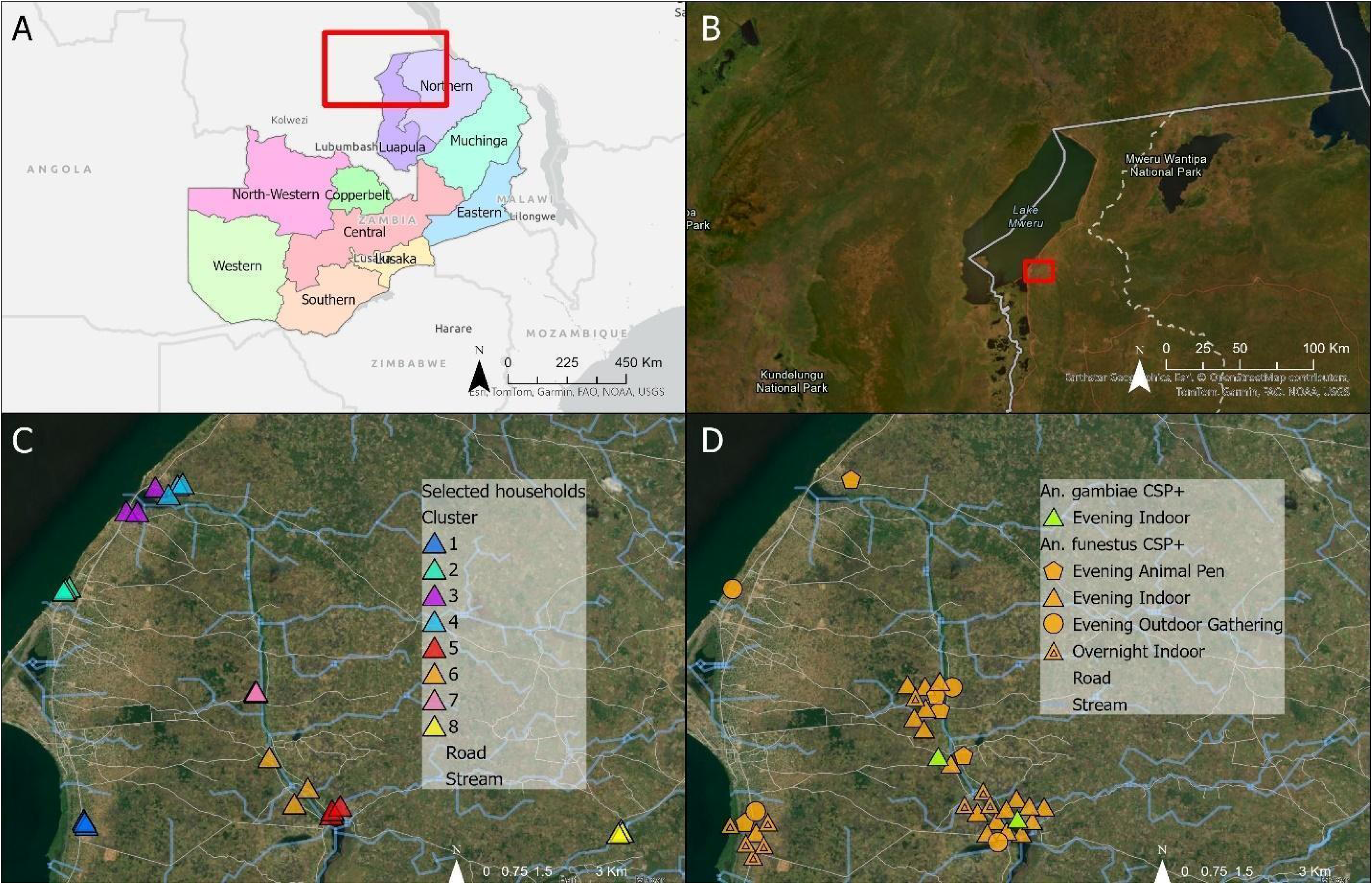
Geographical representation of study site and CSP+ anopheline collections. A. Map of Zambia with extent of panel B outlined in red; B. Close-up view of figure A with extent of panels C and D outlined in red; C. All households selected for study, color denoting sampling cluster; D. All CSP+ anophelines captured in the study, *An. funestus* s.s. in orange and *An. gambiae* s.s. in green. Shape outlines indicate the trap type of the sample.

### Household selection and environmental covariates

Due to established differences in vector intensity between lakeside and inland regions, eight clusters of three households (n = 24 total) were selected for this study from lakeside (n = 12) and inland (n = 12) areas (**Fig 1C**). “Inland” households are defined as households >3 km from the lake. Households were selected using convenience sampling from sites with ongoing ICEMR data collection to reduce logistical constraints. To increase diversity of anophelines from sampled households, half of inland households were selected from areas that received IRS the previous October while the other half did not receive IRS. Eligible households were required to have at least one animal pen outside of the main sleeping structure. At the time of enrollment, a survey regarding household characteristics and human behavior was conducted. Survey questions included employment status and education level of the head of the household, house construction materials, malaria prevention, and animal ownership. This study was approved by the Johns Hopkins Bloomberg School of Public Health (IRB: 00003467) and the Zambian Tropical Diseases Research Centre (IRB: TDRC/ERC/2010/14/11). Household coordinates were collected using a GPS device (Garmin GPSMAP 78sc) and uploaded into ArcGIS Pro (ESRI, Redlands, CA, USA). Streams were previously mapped and classified [21], and distance to the nearest stream was calculated using the “near” tool in ArcGIS Pro.

### Entomological sampling and household surveys

Entomological collections took place 3 nights/week every other week in two of the inland clusters and two lakeside clusters, and 3 nights/week every other week in the remaining four clusters for 6 weeks. At each visit, a single miniature CDC LT (John W. Hock Co., Gainesville, FL, USA) was suspended 1.5-1.8 m off the ground in one of three locations, rotating following a Latin-square design: the sitting room of the sleeping structure, an outdoor area where people gathered in the evening, or next to an animal pen. Placing a single trap at the household on any given night ensured that there was no competition between traps and the Latin-square design ensured that all placements were sampled in each household every week. The head of household was asked to turn the trap on at 16:00 and tie the collection cup at 22:00. In the third replicate, overnight collections were included for indoor traps only, and householders were instructed to replace the trap collection cup with a second empty cup at 22:00 and tie the second cup at 06:00. Outdoor overnight collections could not be carried out due to concerns about equipment security. The study team collected traps and collection bags the next morning and conducted surveys regarding household features and human behavior.

### Mosquito processing

Mosquito specimens were killed by freezing, morphologically identified at the field station in Nchelenge [22], and stored individually on silica gel at room temperature. Desiccated samples were returned to the Johns Hopkins Bloomberg School of Public Health (BSPH), where each specimen was split into head/thorax and abdomen. All specimens from the overnight collections were processed (n=581). However, due to the high counts of *An. funestus* captured in the early-evening collections, a temporal and geographically representative subset was selected (n = 1156/1879; 62%) for molecular processing and identification. Samples were prioritized for inclusion in species verification if they were visually blooded. Abdomens of early-evening specimens were crushed in lysis buffer using a Qiagen Tissue Lyser II instrument (Qiagen, Germany) and DNA was recovered using an automated extraction method with a QIACube HT (Qiagen, Germany) at Purdue University [23]. Abdomens of overnight specimens were manually crushed in lysis buffer using sterilized pestles and DNA was extracted using the Qiagen DNeasy Blood & Tissue Kit (Qiagen, Germany) following the manufacturer’s protocol at BSPH.

Samples morphologically identified as *An. funestus* or *An. gambiae* underwent previously described PCR-based analysis for each species complex [24, 25]. Samples identified as *An. squamosus* underwent amplification through a previously published PCR assay using primers specific to *An. squamosus* [26]. A separate PCR-based assay targeting the nuclear ITS2 region was performed on any specimen that failed any of the previously listed PCR assays or was morphologically identified as a different anopheline species [27]. Samples morphologically identified as *An. gibbinsi* underwent a previously detailed molecular processing scheme for confirmation [28].

Several species, including *An. rufipes, An. maculipalpis,* and *An. pretoriensis*, produce an amplicon size of ∼500 bp from the ITS2 PCR assay. A multiplexed PCR designed to differentiate these three species was developed at the U.S. Centers for Disease Control and Prevention (S1 Table) [Adeline Chan, personal communication]. This assay was applied to any sample that was morphologically identified as *An. maculipalpis* or was unidentified and produced a 500 bp amplicon on the ITS2 assay. Only the ITS2A, RUF-R, and MACU-R primers were included in the master mix for these samples. All indeterminate or negative samples underwent ITS2 and/or cytochrome oxidase I (COI) sequencing [27]. Misassignment was calculated as the percentage of specimens within a molecularly confirmed species that were incorrectly classified by morphology. Misidentification was calculated as the percentage of specimens within a morphologically identified group that were molecularly confirmed to be a different species. All visually blooded *An. funestus* were processed through a PCR assay targeting the vertebrate 12S ribosomal RNA gene that differentiates between human and other vertebrate hosts [29]. These results were used to calculate the human blood index (HBI) by dividing the number of samples with human DNA divided by the number with any detectable vertebrate host DNA.

### Sequencing

Sanger sequencing was performed at the Johns Hopkins Medical Institutions Synthesis and Sequencing Facility on the ITS2 and/or COI gene regions of samples with negative or ambiguous PCR results [27]. Forward and reverse sequences were imported into Geneious Prime version 2021.2.2 (https://www.geneious.com), trimmed to remove low-quality reads, and aligned to create a consensus sequence [26, 30]. If sequencing failed multiple times in one direction, the read from a single direction was used. Any remaining primer sequences were also trimmed from each sequence. Consensus sequences were compared to the NCBI’s GenBank database and reference samples using BLASTn, and final identifications to the species level were confirmed if they had >99% identity to a NCBI GenBank sequence. Sequences were submitted to NCBI’s GenBank, and accession numbers were acquired for both ITS2 (GenBank accession PV972246-PV972336) and COI (PV963682-PV963768).

### Parasite detection

Heads/thoraces were homogenized and circumsporozoite protein (CSP) ELISAs were performed using controls and protocols from BEI Resources Malaria Research and Reference Reagent Resource Center to detect the presence of CSP from *Plasmodium falciparum* sporozoites [16, 30]. Samples were run in duplicate pools of five mosquito homogenates for the first ELISA, and then run individually in duplicate if the pool was positive [26, 31, 32]. CSP-positive samples were verified to have *P. falciparum* DNA by *pf*LDH qPCR [26, 30, 33].

### Human biting rate (HBR), human biting index (HBI), sporozoite infection rate (SIR), and entomological inoculation rate (EIR) calculations

Indoor human biting rates (HBRs) were estimated by dividing the number of captured *An. funestus* or *An. gambiae* by the number of people that reported sleeping in the house the night of collections. Outdoor HBRs were calculated by dividing the number of *An. funestus* or *An. gambiae* by the number of people reported to be near the trap while it was running. Human blood index (HBI) was calculated by dividing the number of specimens positive for human DNA by the total number of specimens with detectable blood meals. Sporozoite infection rate (SIR) was calculated by dividing the number of qPCR-confirmed ELISA-positive *An. funestus* or *An. gambiae* by the total number that underwent ELISA. The entomological inoculation rate (EIR) (the number of infectious bites per person per 6 months) was calculated by multiplying the mean HBR by the SIR. The EIR was calculated for a 6-month period as all sampling was conducted during the dry season and prior data have demonstrated that vector counts, especially those of *An. funestus*, are much lower in the wet season.

### Statistical analyses

Data from anophelines collections and household surveys was analyzed to identify risk factors associated with early-evening *An. funestus* foraging. A univariate negative binomial generalized linear mixed-effects model was fit to the number of *An. funestus* per trap separately for each predictor variable using the glmer.nb function from the MASS package in R. A household-level random intercept term was included to account for repeat visits. Multivariate models were also performed using the glmer.nb function from MASS with a household-level random intercept term. The model of best fit was selected using forward-selection Akaike information criterion (AIC). A paired Wilcoxon rank-sum test was performed to compare the number of *An. funestus* captured indoors in the early-evening compared to overnight.

## Results

### Household information

Each household was visited 9 times, resulting in 216 early-evening collections and 24 overnight collections. Among early-evening traps, six were accidentally turned off after 22:00 and the associated anophelines (n = 21) were excluded, leaving 70 animal pen collections, 69 outdoor gathering collections, and 71 indoor collections remaining for analysis. Overnight collections were successfully performed in all 24 households.

Household construction quality of lakeside houses tended to be better than inland houses, with more lakeside houses featuring brick/concrete walls and metal roofs (33.3%) than inland houses (8.3%) (p = 0.04) (S2 Table). Open eaves were present in 83.3% (n = 10/12) of inland and 75% (n = 9/12) of lakeside houses (S2 Table). The number of rooms (3.5 ± 0.9) and number of occupants (5.7 ± 2.9) were smaller in inland houses compared to lakeside houses (4.9 ± 1.5, 7.1 ± 2.8) (p = 0.01) (S2 Table). Inland households had a higher proportion obtaining water from streams or rivers (16.7%) compared to lakeside households (8.3%), which had a higher proportion of borehole water sources (37.5%) (p = 0.003) (S2 Table). Ownership of goats (45.8%), dogs (25%), chickens (37.5%), and ducks (16.7%) was higher among lakeside versus inland households (37.5%, 4.2%, 33.3%, 4.2%, respectively), but the ownership of pigs (25%) and cats (12.5) was higher among inland houses compared to lakeside houses (16.7%, 4.2%) (S2 Table).

### Spatial and trap type distribution of early-foraging anophelines

More anophelines were captured in and around inland households than lakeside households (Fig 2). The median number of anophelines per trap was highest in indoor traps in both inland (median = 32, IQR = 5-51) and lakeside (median = 1.5, IQR = 0-6.75) locations. Inland, this was followed by animal pens (19, IQR = 8-29.5) and outdoor gathering traps (7, IQR = 3-18). Lakeside, the median number of anophelines per trap in both outdoor gathering traps and animal pens was 0 (IQR 0-2 and IQR 0-1, respectively).

**Fig 2.**
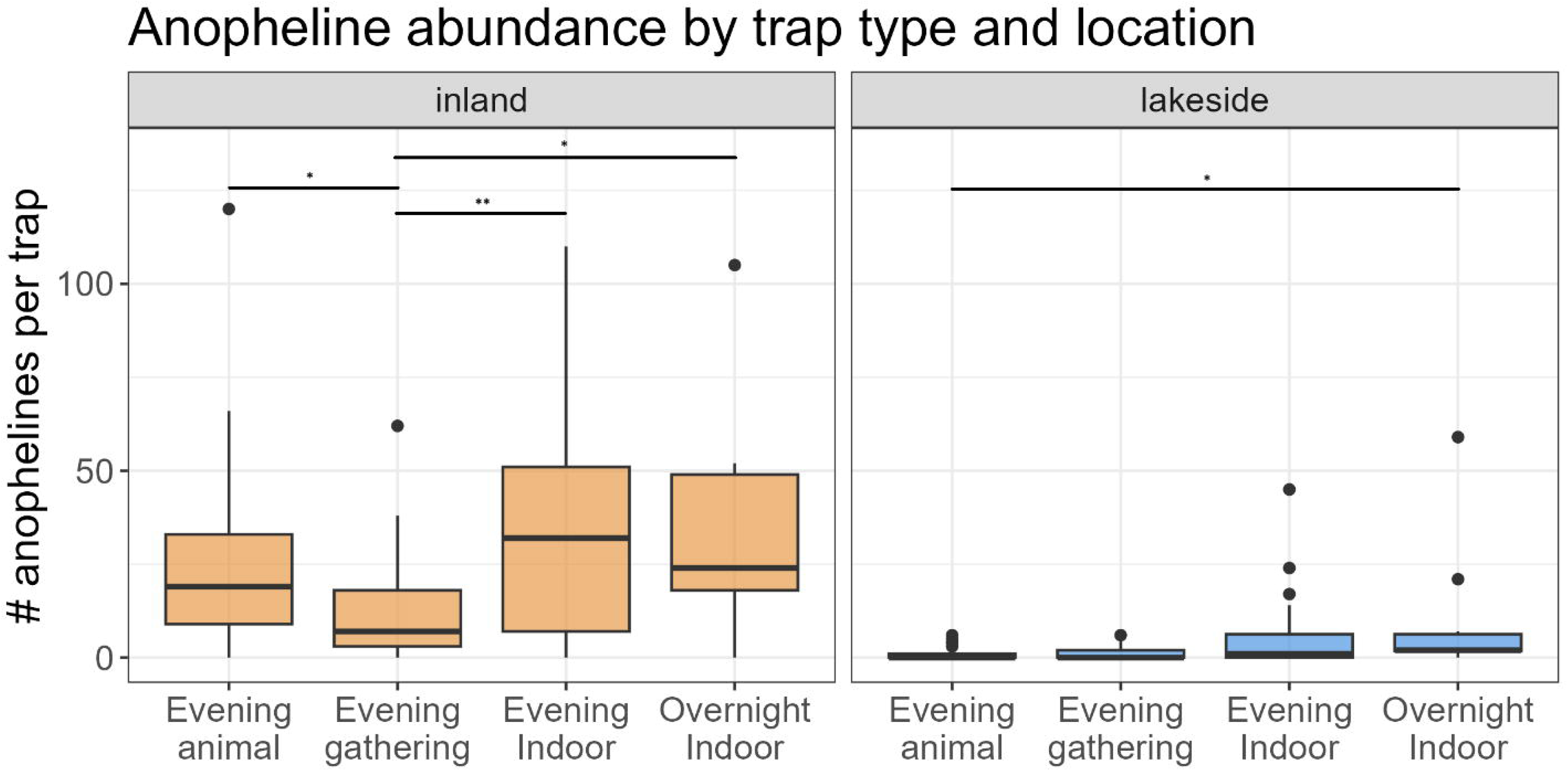
Early-evening anopheline abundance per trap, stratified by trap type and location. * = p<0.01, ** = p<0.005, *** = p<0.001 using pairwise Wilcoxon rank sum test. Unlabeled pairwise comparisons were not significantly different.

### Species composition

Of the 2,734 female early-evening anophelines collected in this study, 1,974 were selected for molecular processing. Those not processed included the early-evening exclusion subset of *An. funestus* (n = 716), samples lost in transit (n = 6), and samples with no abdomen (n = 38). DNA extraction failed for 6/1,974 (0.3%) samples. Final molecular identification for missing samples, samples with no abdomen, or failed extraction samples were recorded as “unidentified”, and *An. funestus* that were not processed were recorded as *An. funestus* sensu latu (s.l.). All overnight female anopheline samples (n = 581) were successfully processed.

Twelve anopheline species were molecularly identified from early evening collections. The majority were *An. funestus* s.s. or *An. funestus* s.l. (n = 1,865, 68.2%), captured primarily in indoor traps (n = 1,370, 73.4%), followed by outdoor gathering traps (n = 285, 15.2%) and animal pen traps (n = 210, 11.2%) (S3 Table). *Anopheles gambiae* s.s., the other recognized vector in Nchelenge, made up only 1.9% (n = 52) of captured anophelines, and was found primarily indoors (n = 41, 66.1%). The other ten *Anopheles* species captured were collected predominantly or exclusively in inland traps, revealing greater diversity compared to lakeside collections (S3 Table)*. Anopheles funestus* also dominated overnight indoor traps, comprising 96.9% of total overnight anophelines (n = 564), while *An. gambiae* made up only 1.7% (n = 10) (S3 Table).

Across all specimens, 1.2% were misidentified and 3.4% were misassigned (S4 Table). Species without defined morphological characteristics (*An. sp. 9, An. sp. 15, An. sp. UG1, An. sp. UG2)* and species with only one specimen (*An. theileri, An. rufipes)* were all misidentified in the initial classification (S4 Table). Misassignment was highest among *An. squamosus* (8.2%, n = 11/134) and *An. maculipalpis* (3.2%, n = 3/90) (S4 Table). *Anopheles funestus, An. gambiae, An. gibbinsi,* and *An. paludis* had < 2% misassignment (S4 Table).

### Anopheles funestus host preference

Of all early-evening *An. funestus* captured with an intact abdomen (n = 1,832), 11.4% were recorded as visually blooded in the field (n = 208) of which 85% were captured indoors (n = 177) (S5 Table). Indoor evening traps were significantly more likely to contain visually blooded *An. funestus* compared to other evening trap types (p < 0.001) (Fig 3). Host preference assays for *An. funestus* and *An. gambiae* demonstrated a 100% HBI for both species. The proportion of visually blooded *An. funestus* was not statistically different in early-evening traps compared to overnight traps run in the third replicate (Fig 3).

**Fig 3.**
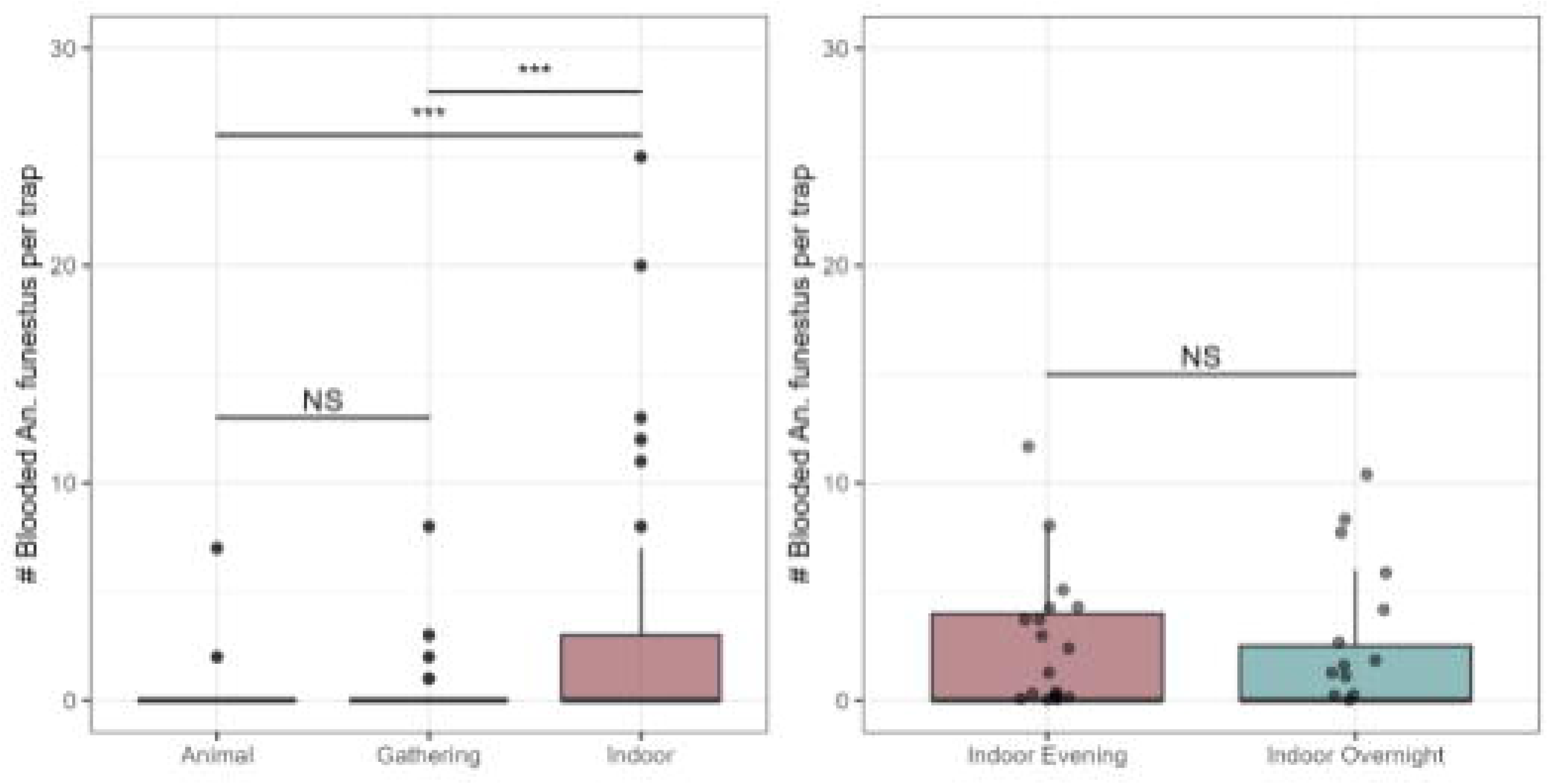
Number of blooded *Anopheles funestus* per trap. Left: Number of blooded *An. funestus* per trap from evening collections (16:00-22:00), stratified by trap type and location. Right: Number of blooded *An. funestus* per trap from third replicate only, comparing early indoor evening and overnight traps. Three asterisks indicate p<0.001 and NS = not significant using pairwise Wilcoxon rank sum test.

### CSP ELISA results

Homogenization and CSP ELISAs were performed on 2,725/2,734 (99.7%) early-evening female anopheline heads/thoraces. *Anopheles funestus* s.s. and *An. gambiae* s.s. were the only species positive for *P. falciparum-*CSP by ELISA. Every specimen positive by CSP ELISA was also confirmed positive using the *PfLDH* qPCR. Proportionally, positivity among evening catches was higher among *An. gambiae* compared to *An. funestus* (3.7% vs. 1.5%). All *P. falciparum*-positive *An. gambiae* were captured in inland, indoor traps. Five of 27 (18.5%) early-evening *P. falciparum*-positive *An. funestus* were captured in lakeside households. Of those, two (40%) were captured near animal pens, two (40%) were captured in outdoor gathering areas, and one (20%) was captured indoors (Fig 1D). Among the *P. falciparum*-positive *An. funestus* captured inland, 17 (77.3%) were captured indoors, three (13.6%) were captured near animal pens, and two (9.1%) were captured in outdoor gathering areas (Fig 1D). Only *An. funestus* was found positive for *P. falciparum*-CSP from indoor overnight collections. Four out of 458 (0.87%) were positive from inland traps, while 5/97 (5.17%) were positive from lakeside traps (Fig 1D).

### HBR, SIR, and EIR calculations

HBR, SIR, and EIR for anophelines were calculated stratifying by three levels (species, trap type, and location). The highest calculated EIR in the study was from *An. funestus* in early-evening inland indoor traps (Table 1). Estimated EIRs between 16:00-22:00 for *An. funestus* ranged from 1.3 infectious bites/person/6 months (ib/p/6mo) in lakeside gathering traps to 18.8 ib/p/6mo in inland indoor traps (Table 1). The total estimated EIR for early-foraging *An. funestus* was 12.3 ib/p/6mo. The only estimated EIR above 1 for *An. gambiae* was among inland, indoor traps (Table 1). EIRs were not calculated for animal pen traps because data regarding potential human contact was not collected. EIR estimates for overnight indoor foraging (22:00-06:00) *An. funestus* (16.0 ib/p/6mo) were similar to the early-evening indoor EIR (12.3 ib/p/mo), leading to an estimated 28.3 total ib/p/6mo (Table 1).

**Table 1.** EIR calculations.

| Species | Location | Trap type | N traps | Mean HBR | SIR (%) | EIR | EIR 95%CI |
| --- | --- | --- | --- | --- | --- | --- | --- |
| *An. funestus*<br>early evening | Inland | Indoor | 36 | 7.61 | 1.35 | 18.75 | (12.30-25.19) |
|  |  | Gathering | 34 | 1.27 | 1.35 | 3.13 | (1.58-4.66) |
|  | Lakeside | Indoor | 34 | 1.44 | 2.20 | 5.78 | (2.59-8.95) |
|  |  | Gathering | 32 | 0.33 | 2.20 | 1.32 | (0.26-2.42) |
|  | Total | Indoor | 70 | 4.61 | 1.46 | 12.28 | (8.11-16.47) |
|  |  | Gathering | 66 | 0.81 | 1.46 | 2.15 | (1.20-3.14) |
| *An. gambiae*<br>early evening | Inland | Indoor | 36 | 0.15 | 4.26 | 1.17 | (0.61-1.79) |
|  |  | Gathering | 34 | 0.07 | 4.26 | 0.54 | (0.04-1.02) |
|  | Lakeside | Indoor | 34 | 0.03 | 0 | 0 | NA |
|  |  | Gathering | 32 | 0.01 | 0 | 0 | NA |
|  | Total | Indoor | 70 | 0.09 | 3.70 | 0.61 | (0.33-0.93) |
|  |  | Gathering | 66 | 0.04 | 3.70 | 0.27 | (0.03-0.48) |
| *An. funestus*<br>overnight | Inland | Indoor | 12 | 9.50 | 0.87 | 15.08 | (1.99-28.17) |
|  | Lakeside | Indoor | 12 | 1.44 | 5.15 | 18.74 | (0-28.46) |
|  | Total | Indoor | 24 | 5.47 | 1.60 | 15.97 | (3.05-28.88) |
| *An. gambiae*<br>overnight | Inland | Indoor | 12 | 0.19 | 0 | 0 | NA |
|  | Lakeside | Indoor | 12 | 0 | 0 | 0 | NA |

|  | Total | Indoor | 24 | 0.09 | 0 | 0 | NA |
| --- | --- | --- | --- | --- | --- | --- | --- |

EIR stratified by species, trap type, and location for early evening mosquitoes. HBR = human biting rate (bites/person/night); SIR = sporozoite infection rate; EIR = entomological inoculation rate (infectious bites per person per 6 months).

Estimations of SIR and HBR that did not stratify by site and location missed important patterns in the EIR for both *An. gambiae* and *An. funestus.* For example, calculating the early-evening EIR for all indoor early-biting *An. funestus* provided an overall estimate of 12.3 ib/p/6mo, but after stratifying by location the EIR was 18.8 ib/p/mo in inland households and 5.8 ib/p/6mo lakeside.

### Risk factor analysis

In the univariate analysis, household level risk factors significantly associated with a higher abundance of early-foraging *An. funestus* were inland and indoor locations, open wells, natural wall materials, open eaves, and coal and charcoal cooking materials (S6 Table). An increased number of rooms in a household and burning a fire within 5 m of the trap were associated with decreased abundance of *An. funestus* (S6 Table). Many of these variables correlated with each other. For example, inland households were more likely than lakeside households to have fewer rooms, natural wall materials, and open wells as a water source. The multivariate model with best fit contained only household location and trap location. The strength of the positive association between *An. funestus* abundance and indoor traps increased after controlling for location (S6 Table).

### Human activity

Among lakeside households, 83% reported the last household member was indoors and 64% were asleep before 22:00 (Fig 4). Among inland households, 90% reported the last household member was indoors and 67% were asleep before 22:00. 60% and 64% of households reported the first member leaving by 06:00 in lakeside and inland areas, respectively (Fig 4). Inland and lakeside households reported a similar proportion of ITN use (56% and 54%) (Fig 4).

**Fig 4.**
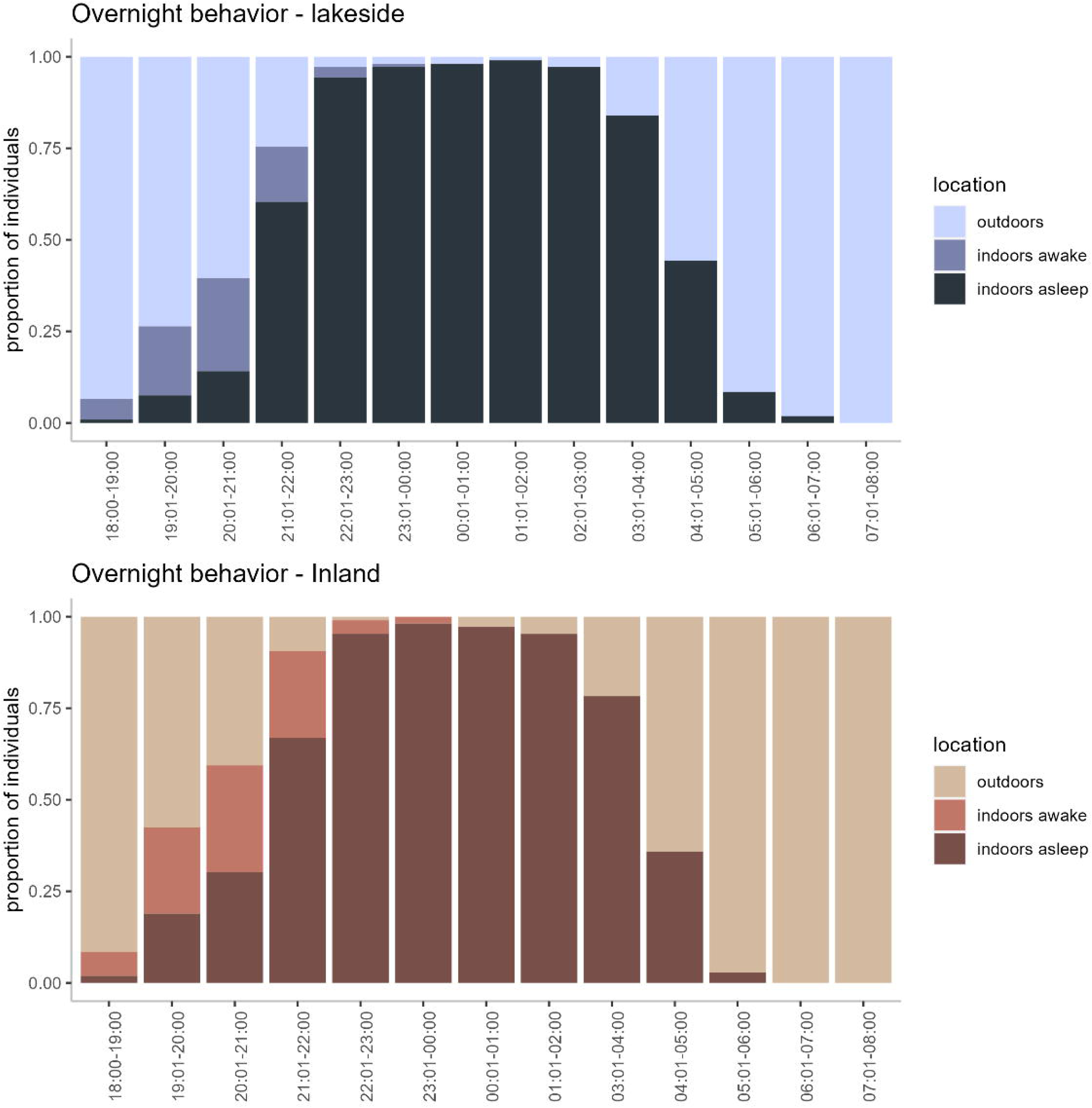
Overnight human behavior. Proportional time spent by participants throughout the evening, stratified by location in study area.

## Discussion

This study explored the hypothesis that the intransigence of malaria in Nchelenge District is due in part to early evening and outdoor foraging of local anophelines. Twelve different species of anophelines were collected by early-evening CDC LTs over a 2-month period during the dry season in 2019. Seventy-percent of these specimens were *An. funestus,* which were captured primarily indoors but also in outdoor gathering areas and near animal pens. Human blood was found in specimens from all three trap locations, suggesting that this major vector in Nchelenge District is active and likely host seeking in and around homes through the early evening hours before 22:00. However, vector abundance was spatially heterogeneous. Traps set at inland households captured 92% of all early-evening anophelines, indicating a higher risk of vector exposure in inland areas, consistent with previous findings [1, 2, 34].

Other anopheline species made up 30% of collections, some of which have been previously implicated in malaria transmission across Africa [26, 31, 35–37]. Given their tendencies to forage outdoors, these other species are likely to easily evade indoor-targeted malaria control tools. Although none of the non-*An. funestus*, non-*An. gambiae* species were positive for *P. falciparum* sporozoites in this sample set, parasite prevalence among humans is extremely high, with RDT rates reaching 40-63% in some regions of Zambia [38], and it is likely that even occasional human blood meals will lead to ingestion of parasites. Future studies should continue to monitor these species for transmission potential by assessing their sporozoite rates, human blood index and biological capacity to support *Plasmodium* parasites.

Two species of anophelines were positive for *P. falciparum* in this study, the primary vectors; *An. funestus* and *An. gambiae.* With an overall CSP positivity of 1.5%, CSP-positive *An. funestus* were captured at all three early-foraging trap locations, suggesting that people have the potential to encounter infectious vectors indoors and outdoors before 22:00. However, risk varied spatially by both location in the study area and within a household, which was further demonstrated in the results from the univariate and multivariate analyses. Inland and indoor traps were consistently associated with higher *An. funestus* abundance. This study tracks with a previous analysis of anopheline diversity in Nchelenge with an estimated 1.2% infection rate in *An. funestus* and significantly greater abundance of this vector in inland households [34].

Typically, EIRs for a certain species are reported as a single number across a study area, which may not encapsulate the localization of vector foraging activity as EIRs have been found to vary widely even within a village driven by ecological, spatial, and temporal factors [39]. Spatial differences in EIR in this study were driven by low HBR at lakeside households compared to inland households. The indoor *An. funestus* early-evening EIR is estimated at 18.8 ib/p/6mo in inland households while only 5.8 ib/p/6mo in lakeside households. If the *An. funestus* EIR is reported as a single number, the spatial resolution of risk is lost and a single number of 12.28 ib/p/6mo is reported, overestimating risk at lakeside households and underestimating risk at inland households. Likewise, future studies should aim to assess HBR and EIR outdoors as *An. funestus*, typically considered an endophagic vector, has exhibited greater HBRs outdoors compared to indoors in other regions [40].

When comparing the early-evening and overnight collections from the third replicate, there were no differences in anopheline abundance, SIR, or proportion blooded, suggesting that there may be similar risk before and after 22:00. However, differences in early-evening and overnight mosquito activity in and around households that we interpret as ‘foraging’ cannot be discerned by CDC light trap collections. Studies in western Kenya, Tanzania, and Mozambique have suggested a similar trend in which malaria vectors have initiated foraging or feeding behaviors earlier in the evening than previously accepted while people are awake and unprotected by nighttime and/or indoor focused interventions [41, 42, 43]. Interestingly, *An. funestus* began biting early in the evening in the western Kenya study, but displayed peak biting during 05:00-06:00, further challenging the widely described “middle-of-the-night” behavior of this species [42]. Future human landing catch studies will provide further insight into the temporal host-seeking behavior of these anophelines. This study also demonstrated that many people are still awake and outdoors while these early-evening vectors are active. Even after going to sleep, only 56% of people inland and 54% of lakeside residents reported using an ITN, leaving many individuals with no protection against host-seeking malaria vectors at any point during the day or night if IRS was not applied to the structure or if the IRS has decayed since application.

Misassignment and misidentification were lower in this study than other studies reported from Africa [27, 44, 45]. Misidentification was lowest among species commonly collected in high numbers, followed by species with distinctive features on the abdomen or wing. Higher misidentification rates were likely due to their morphological similarities among species and reliance on defining characteristics on the wing and hind tarsi, which are often damaged during specimen collection and transport. The difficulty in morphological classification highlights the need for additional molecular tools for identification verification as some of these taxa are significant malaria vectors at other localities in sub-Saharan Africa, yet may vary morphologically by geographic origin even within a singular species [46–47].

There were several limitations to this study. First, the convenience sampling strategy was not random and therefore may not reflect the entirety of the district, and the short period of study may not be representative of other seasons. Second, CDC LTs may not accurately represent foraging and biting times as well as other metrics like human landing catches. Efforts should be made to expand sampling of early-foraging anophelines both spatially and temporally, using other methods to quantify specific biting times and rates of these species. Third, human behavior was only assessed through a survey asking when the last person entered, went to sleep, and left the house in the morning. The design of the study (asking householders to turn off the trap) could have altered the behavior of the participants, and it also does not account for the behavior of other household members. Finally, HBR and EIR calculations used the number of people sleeping in the household as a denominator, assuming all people in the household are equally exposed to host-seeking vectors. Some household members may be less attractive to mosquitoes or enter and exit the house multiple times, and mosquitoes may not be continuously foraging at the same rate throughout the entire evening.

Overall, *An. funestus* and *An. gambiae* remain the major vectors in Nchelenge District during the dry season and must continue to be targeted for control efforts successful malaria control. In this study, most of these mosquitoes were captured indoors and should be susceptible to indoor interventions. However, this study also provided evidence of possible behavioral resistance to indoor interventions, including outdoor and early-evening foraging, and other anopheline species with exophagic behaviors. Human activity indoors before sleeping and outdoors allows for the potential of transmission before sleeping. This study also highlighted the importance of accounting for the degree of spatial heterogeneity when considering EIRs. Overall, this study adds to the growing body of evidence that malaria transmission is not limited to late-night, indoor settings in Nchelenge District, Zambia.

## Data Availability

Under the National Health Research Act, the Government of Zambia does not allow public access to data collected in Zambia. All investigators interested in the datasets supporting the conclusions of this article are required to submit a written request to the Ministry of Health. Contact the TDRC IRB Chairperson ( +260212 615444) to coordinate the request.

## Acknowledgements

The authors gratefully acknowledge the Southern Africa ICEMR field teams in Nchelenge and the Tropical Diseases Research Centre (TDRC) for their logistical support and participation in field collections. The authors are also very grateful to the communities in Nchelenge District, Zambia, for their participation with this study. The authors would also like to thank Jack Dorman, Dr. Ilinca Cuibotariu, and Dr. Giovanna Carpi at Purdue University for performing the abdominal DNA extractions. The protocols and primers for detecting *Anopheles pretoriensis/ rufipes/ maculipalpis* were developed by Drs. Lijuan Wang and Adeline Chan at the U.S. Centers for Disease Control and Prevention. The reagents for the CSP ELISAs were obtained through BEI Resources, National Institute of Allergy and Infectious Diseases, National Institutes of Health. The *Plasmodium falciparum* Sporozoite ELISA Reagent Kit, MRA-890, was contributed by Robert A. Wirtz.

This study was supported in part by funding from the NIH Southern and Central Africa International Centers of Excellence for Malaria Research (U19AI089680), Johns Hopkins Malaria Research Institute, Bloomberg Philanthropies, and NIH T32 Training Grants (T32 AI007417 & T32 AI138953).

## Supporting Information

**S1 Table. Conditions for maculipalpis/pretoriensis/rufipes species-specific PCR.** This PCR uses the shared ITS2-A forward primer with species-specific reverse primers (RUF, 261 bp; MACU-R, 420 bp; and PRET-R, 422 bp). Each 25 µL PCR reaction consisted of 1x buffer, 1.0 mM dNTPs, 2 U of *Taq* polymerase, 50 pmol of each primer, and 1.0 µL extracted abdominal DNA. MultiGene OptiMax thermal cycler (Labnet International, Inc., Edison, New Jersey, USA) conditions consisted of an initial denaturation of 2 minutes at 95°C, followed by 35 cycles at 94°C for 30 seconds, 50°C for 30 seconds, and 72°C for 40 seconds. The final extension step was 72°C for 5 minutes.

**S2 Table. Household information.**

* Household was visited by a test and treat program within the last year

+ Household was visited by a filariasis MDA campaign within the last year

**S3 Table. Molecular identification of species by trap type and location.**

*Overnight collections were only performed indoors for one replicate.

**S4 Table. Morphological vs. molecular assignment of all molecularly processed specimens.**

*n includes all samples that were molecularly confirmed (excludes 716 An. funestus samples that were not processed)

+misidentification calculation excludes samples that were morphologically unidentified misid = misidentified; misassign. = missassigned

**S5 Table. Visual *An. funestus* blooded abdomen status by trap type.**

* Excluded specimens missing an abdomen

+ Indoor overnight trap specimens were only collected for one replicate

**S6 Table. Household level risk factors associated with higher abundance of *Anopheles funestus*.** Univariate and multivariate results from negative binomial generalized linear mixed-effects model performed with # *An. funestus* per trap as the outcome.

## Notes

### Competing Interest Statement

The authors have declared no competing interest.

### Author Declarations

The study was approved by the Zambian Tropical Diseases Research Center (TDRC) under IRB no: TDRC/C4/09/2018 and the Johns Hopkins Bloomberg School of Public Health (Baltimore, Maryland) under IRB no: 00003467.

